# Stressors, Manifestations and Course of COVID-19 Related Distress Among Nurses and Midwives in Tasmania

**DOI:** 10.1101/2021.11.29.21266774

**Authors:** Kathryn M. Marsden, Julie M. Porter, IK. Robertson

## Abstract

The deleterious effects relating to the COVID-19 pandemic on the mental health of healthcare workers has now been widely established. Understanding how COVID-19 affects their work and life is complex and multidimensional. This study describes the critical stressors and how they manifest within both the work and larger social environment for nurses and midwives in Tasmania, Australia.

A longitudinal, descriptive survey was designed to explore the trajectory of the psychological health of Tasmanian public sector nurses and midwives during the COIVD-19 pandemic. The survey was distributed at 3 timepoints over a 12-month period and consisted of a battery of psychological tests which included the Patient Health Questionnaire, General Anxiety Disorder, Insomnia Severity Index, and the Impact of Events Scale-Revised, together with free text comments.

The associations between outcome and predictor variables were assessed using mixed effects linear regression and linear mixed model analyses. Free text comments were themed.

High levels of stress and mental exhaustion were attributed to threatened workplace team culture; compromised quality of patient care; the impact on family, home, financial and economic domains; lack of clear communication; issues surrounding personal protective equipment; and female gender. Study data show younger nurses and midwives suffered higher levels of stress and mental exhaustion than older.

This study highlights the need for stable and functional relationships at home and at work for nurses and midwives.

Factors which will help preserve the mental health of nurses and midwives include strong workplace culture with ongoing processes to monitor organisational burnout; building resilience, particularly among younger nurses and midwives; protection of healthcare worker safety; clear communication processes and supporting stable and functional relationships at home. The health service has an imperative to ensure optimum service delivery by safeguarding staff, despite the inevitable health stress imposed by the nature of the work.

## INTRODUCTION

The deleterious effects relating to the COVID-19 pandemic on the mental health of nurses has now been widely established. An abundance of literature consistently reports the increased prevalence and severity of depression, anxiety, stress, sleep disturbance among nurses in Australia (1-6) and globally (7, 8). Understanding how COVID-19 affects a nurse’s work and life is complex and multidimensional. This longitudinal study describes the stressors and how they manifest in the context of the larger social environment. This will assist the design of comprehensive support and prevention strategies to reduce psychological distress for nurses and midwives in Australia.

International research data relating to COVID-19 and healthcare workers (7, 8) show one third of nurses have experienced greater psychological distress, evidenced by level of nervousness, agitation, psychological fatigue and depression in the past four weeks compared with the general population (9) and are at high risk for developing stress response syndromes, post-traumatic stress disorder, chronic illness, and burnout (10).

Maqbali et. al. noted in their systematic review and meta-analysis of over 93,000 nurses, summary prevalence of stress, anxiety, depression, and sleep disturbance among participants at 43%, 37%, 35%, and 43% respectively (7). Publications describing high psychological distress among healthcare workers predominately originate from countries with high COVID-19 prevalence, directly associating high risk of infection from patients to high risk of psychological distress Authors acknowledge a longitudinal study is needed to distinguish psychological symptoms during and after the infectious disease outbreaks and the design of comprehensive support strategies to reduce psychological distress (7, 11).

Lessons learned from previous epidemics help us understand reactions to COVID-19. Previous studies on Severe Acute Respiratory Syndrome (SARS) and Ebola uncover the serious nature of psychological distress endured by healthcare workers, revealing the long-standing nature of the distress with many nurses suffering post-traumatic stress disorder (PTSD), depression, anxiety, and burnout persist even after the outbreaks (12, 13). Research experiences of health service staff during the SARS outbreak in 2003 identified additional factors including quarantine, job stress, poor organisational support, and stigmatisation which all impacted on nurses’ psychological distress (14-16). We are learning that psychological distress from COVID-19 and natural disasters (17) have long-standing effects in the general population (18) and will not merely snap-back to a pre-COVID state of life. From experience with bushfires and trauma, it is often a few months down the track when psychological issues come to the fore (17). As circumstances ease and communities once again adopt freer living conditions, there may still be residual psychological distress experienced by many. Acute psychological distress can progress to a chronic state and for healthcare workers this can precipitate burnout(19). Unlike Coronaphobia (20), or Covid Stress Disorder, considered a type of adjustment disorder which subsides once the pandemic subsides, the COVID-19 Stress Syndrome persists (11).

The Australian COVID-19 experience has been mild in comparison to the international scene, with relatively low case numbers, mortality rates and hospitalisations. Still, Australian population data related to COVID-19 uncovers significantly greater negative emotions among nurses compared to Australian norms (21, 22). Published data relating to the mental health effects of COVID-19 among Australian nurses and healthcare workers (1-3) shows significant symptoms ranging from mild to extremely severe levels of depression, anxiety, PTSD and sleep disturbance. Research has confirmed from SARS-Cov-1 and Ebola that nurses are vulnerable to psychosocial harm from the COVID-19 pandemic, regardless of the volume of cases or deaths (23-25).

As the pandemic draws out it is allowing us time to adapt. The dynamic nature of the COVID-19 pandemic has necessitated a process of ongoing strategic review to keep abreast with the ever-changing course of the pandemic. New clusters and outbreaks and viral variants have created constant change to daily life heightening pandemic-related stress. Lessons continue to be learned as the pandemic unfolds; we are continuing to build capacity in preparedness (26, 27), such as Health Emergency Management protocols, maintaining substantial stocks of personal protective equipment (PPE), design of comprehensive respiratory protection for broader staff safety and the provision of adequately trained staff to create surge capacity in Intensive Care Units (28). All areas initially proved challenging, creating a lot of perceived risk, stress and negative emotion among staff.

As the pandemic draws out, stress on nurses builds. Nurses have always worked under intense psychological pressure day-to-day (29), long before the COVID-19 pandemic, yet little emphasis has ever been placed on understanding their wellbeing or psychosocial coping mechanisms. Even in the ‘lucky country’ the COVID-19 pandemic remains a global health emergency where nurses provide vital care; pivotal to emergency preparedness and the overall response capacity (29). The longevity of the pandemic is unknown, causing uncertainty and ongoing disruption (30). The additional, acute demands now being placed on nurses due to the current pandemic will arguably add to nurses’ existing distress (31), compounding their risk of stress responses, post-traumatic stress disorder, anxiety, depression, chronic illness, and burnout (27). Burnout has been associated with increased risk of medical errors and poor clinical decision making and may manifest as psychological distress, negative emotions, cause dependence on negative coping mechanisms, and has the potential to exacerbate existing mental unwellness (29).

Understanding fears relating to the pandemic is anything but simple. Overarching factors which precipitate COVID-19 related stress in the general population relate primarily to direct increases in COVID-19 (20). For health professionals, the complexities associated with management of COVID-19 pandemic are multidimensional and each nuance deserves recognition and discussion. COVID-19 impacts immediate and distant home and family, work and employment networks such as conflicting loyalties about commitment to safety for self and risk of viral transmission to family members and commitment to the profession and their employer (29). COVID-19 challenges contamination fears, fears of adverse local and global economic consequences, xenophobia (18) and personal belief systems.

The psychological health of nurses can directly impact patient care. The psychological health of nurses during the COVID-19 pandemic can impact their performance and reduce the quality of patient care. Not being able to provide evidenced-based care, missed care or not being able to maintain standards of care, can further predispose nurses to ethical stress and burnout, compounding the impact on patient care (32). Retaining and supporting the nursing work force requires a focus on promoting and protecting their physical, mental and spiritual health (29).

Healthcare workers are also subject to challenges and changes to daily life. Lockdowns and social restrictions to ‘flatten the curve’ or reduce direct COVID-19 exposure and transmission have also been endured by nurses. Lifestyle changes such as lack of social, leisure and recreation activities impact on physical, mental and spiritual health and can precipitate psychological distress (29, 33, 34). The dynamic effect of isolation and varying stages or levels of fear and vulnerability and its fluctuation over time can overlie or augment precipitating stress factors. Risk factors for psychological distress within the general population include having an underlying illness or disease, living in rural areas, female sex, being at risk of contact with COVID-19 patients and in addition, being a health care worker (8, 11).

Surprisingly while Australia has seen low community prevalence, hospital care workers are shown to have nearly 3 times the risk of infection (35). Tasmania led the way with hospital outbreaks. The rural North West region of Tasmania was unique in that it experienced the first major hospital-based outbreak of COVID-19 in Australia. This nosocomial outbreak occurred during March 2020 (36), at a very early stage of the pandemic which was labelled by the media as the “Epicentre of infection” (37). Local newspaper headlines at the time questioned health workforce integrity indicating that “Misconduct of people impacts on disease transmission”(36). While there was praise of health care workers in the Northern and Southern regions of Tasmania, fear and stigmatisation was instilled amongst the health care workforce in the North West. This left health care workers at the centre of the COVID-19 outbreak feeling a sense of fear, helplessness and blame for their community (36).

Since then, several hospital-based outbreaks have occurred elsewhere in Australia, including the Royal Melbourne Hospital, a major metropolitan hospital. This occurred despite considerable planning, preparation, and resourcing to prevent the nosocomial transmission, escalating the threat and fear among nurses and throughout the entire Tasmanian community. An independent review of this incident in North West Tasmania presented key findings and directions in the areas of protection and prevention for the health workforce to help future-proof the health system for future pandemics (26). Therefore, it is important to examine the Tasmanian nurses and midwives COVID-19 story.

## METHODS

### Study design and participants

This was an on-line, longitudinal, self-reporting, descriptive survey exploring the trajectory of the psychological health of public sector nurses and midwives during the COIVD-19 pandemic.

Following ethics approval, the study was emailed to all public sector nurses and midwives in Tasmania using the Tasmanian Government staff email. Information sheets were attached to the survey as well as distributed to all clinical areas. Posters were erected across health facilities advertising the study. Completion of the online surveys implied consent. The survey was anonymous with respondents entering their own personal identifier on each survey for longitudinal analyses. Survey completion took approximately 20 minutes.

Survey timepoints:

Survey 1: initial survey (April 2020)

Survey 2: 3 months (July 2020) and

Survey 3: 12 months (April 2021).

Each survey was closed out after 3 weeks. Of note is that survey 2 was not circulated to nurses in the North West region due to the critical nature of the COVID-19 hospital outbreak at that time, when all communications were limited to just essential communique.

### Data variables

Data variables included a broad set of demographic and other relevant predictor variables, as below:

∘ Sociodemographic variables. The sociodemographic variables include age, sex, education level, social situation, smoking status, employee classification
∘ Exposure to COVID-19, COVID-19 status, work exposure, community exposure
∘ Workplace support. Access to PPE, COVID-19 testing, access to information and communications, preparation for deployment, workplace team support, workplace support (food, accommodation, and transportation)
∘ Home and family general and financial stress.
∘ Self-perceived health status
∘ Free text comments

Likert scales were used to classify these predictors, and these scales were rank ordered in nature.

### Primary and Secondary Outcome Measures

The primary outcome was self-reported levels of psychological distress (symptoms of anxiety, depression, general stress / PTSD and insomnia) experienced during the 2 weeks prior to the survey.

### Mental Health Assessment Battery

∘ Patient Health Questionnaire (PHQ-9)
∘ General Anxiety Disorder (GAD-7)
∘ Insomnia Severity Index (ISI)
∘ Impact of Events Scale-Revised (IES-R)

#### Patient Health Questionnaire (PHQ-9)

The PHQ-9 is a 9-item measure of depression based on the diagnose criteria of DSM IV. The PHQ-9 has a dual-purpose to screen for the presence of a depressive disorder as well as to grade depressive symptom severity.

The PHQ-9 score ranges from 0 to 27, based on the 9 items scored from 0 = “not at all” to 3 = “nearly every day”. The suggested cut-points are 5, 10, 15, and 20, which represent the thresholds for mild, moderate, moderately severe, and severe depression, respectively (38). Depression severity: 0-4 none, 5-9 mild, 10-14 moderate, 15-19 moderately severe, 20-27 severe. PHQ-9 shows adequate internal consistency at pre- and post-treatment, α = 0.74 and 0.81. The internal consistency for the PHQ-9 based on the current sample was α = 0.77.

#### General Anxiety Disorder (GAD-7)

The GAD-7 is a 7-item measure created as a screening tool for general anxiety disorder and has been used across various settings and populations (39-41) and has a range from 0-21. Anxiety severity ranges: 0-4 none to minimal, 5-7 mild (recommended to monitor symptoms), 8-9 mild though likely to be diagnosed with an anxiety disorder, 10-14 moderate symptoms are clinically significant, 15-21 severe symptoms warrant active treatment. A cut-off score of 10 has been identified as the optimal point for sensitivity (89%) and specificity (82%) (39). GAD-7 has demonstrated strong psychometric properties in the general population and adequate internal consistency across subgroups (α = 0.89). The internal consistency of the GAD-7 based on the current sample was α = 0.89.

#### Insomnia Severity Index (ISI)

The ISI (42) consists of 7 items to assesses the nature, severity, and impact of insomnia. Items include severity of sleep onset, sleep maintenance, early morning awakening problems, sleep dissatisfaction, interference of sleep difficulties with daytime functioning, noticeability of sleep problems by others, and distress caused by the sleep difficulties. A 5-point rating scale is used to rate each item, with 0 = no problem and 4 = very severe problem, yielding a total score ranging from 0 to 28. The total score is interpreted as follows: absence of insomnia (0–7), sub-threshold insomnia (8–14), moderate insomnia (15–21), and severe insomnia (22–28) (43). The ISI has been evaluated in a population-based sample and the internal consistency was excellent, α = 0.90. It is suggested a cut-off score of 10 (86.1% sensitivity and 87.7% specificity) for detecting insomnia in a general population (44). The internal consistency for the ISI-7 in the present study was α = 0.90.

#### Impact of Events Scale-Revised (IES-R)

PTSD often coexists and interacts with anxiety and depression, and contributes to delayed recovery, suboptimal functional outcome, poor quality of life, sleep disorders and feelings of being detached. The IES-R provides a dimensional assessment of PTSD. Participants specify the frequency with which they have had intrusion-, avoidance-, and hyperarousal-related thoughts in the previous 7 days on a Likert scale. The scores for the intrusion component of the scale range from 0-24, for the avoidance component, 0-32 and hyperarousal 0-24 with a total score between 0-88. The higher the score, the greater the level of distress indicated. For the identification of coping disorders, scores above the cut-off point of 35 were classified as a high level of PTSD-related symptoms (45).

Weiss and Marmar (46) report the IES-R showed high internal consistency, with coefficient alphas ranging from 0.87-0.92 for intrusion, 0.84-0.85 for avoidance and 0.79-0.90 for hyperarousal. Test-retest correlation coefficients ranged from 0.57-0.94 for intrusion, 0.51-0.89 for avoidance and 0.59-0.92 for hyperarousal. The IES-R has been used in numerous studies with a wide variety of adult populations and has proved valuable in documenting the course of posttraumatic phenomena over time (47).

## STATISTICAL ANALYSIS

### Quantitative components of analysis

Being a descriptive study, *a priori* calculation of the sample size required for attaining statistical significance was not undertaken.

The four primary outcome scales derived from the battery of psychological tests (PHQ9, GAD7, ISI, IESR 22) are rank ordered with relatively broad ranges but are usually treated as continuous interval variables for purposes of statistical analysis.

The associations between outcome and predictor variables were assessed with mixed effects linear regression and mixed effects ordered logistic regression (linear mixed models), representing “parametric” and the equivalent “non-parametric” analyses to account for the non-interval nature of the measurements. Where it was possible to identify repeated survey completions by the same respondent, this was done, although such identification may have been incomplete.

The mean outcome scores were estimated using mixed effects linear regression, to describe the mean values in the different reported subgroups. Predictor variables were presented as z-scores or standardised normal transformation: ((2)/standard deviation). This allows direct comparison of ORs of each predictor. The relative impact of different predictors on the outcome scores was estimated as odds ratios using multivariate mixed effects ordered logistic regression. An “odds ratio for trend” was estimated in each model representing a way of describing the relative strength and direction of association between two rank-ordered, non-interval measures. All the listed predictors were included in the model for each separate outcome score. The nurse group and sampling time were treated as fixed effects whilst individual effects were assumed to be random.

Missing data for predictor variables were replaced by mean values (a z-score of zero). Missing data for outcome variables was not substituted. Thus, all respondents who completed the outcome questions were included in the regression models, whether or not they completed all the predictor variable questions. All quantitative analyses were performed using Stata MP2 version 16.1 (StataCorp LLC, College Station, Tx USA).

### Qualitative components of analysis

Thematic analysis was used to analyse participant narrative’s looking for patterns, meanings, and potential points of interest at the end of each survey. Thematic analysis was used to derive understanding directly from the participant data. Narrative thematic analysis has been used in conjunction with study data to increase methodological rigour. This person-centred approach compliments the inventory of standardised criteria and outcome severity.

A deductive thematic approach to analysis was used to ascertain themes from the participant narratives, looking for patterns, meaning and potential points of interest in the data, while constantly referencing associated raw data.

## RESULTS

This longitudinal study distributed surveys via email at 3 timepoints during the first 12 months of the COVID-19 pandemic in Australia. Survey 1 n=725, survey 2 n=634 and survey 3, n=456. The dataset was reduced from an apparent 1815 cases to 1676 cases (S1 684; S2 553; S3 439) due to apparent duplicates (survey completions at the same survey time-points by the same respondent, with the exclusion of the second duplicate)

### Demographics

Approximately 90% of respondents across all surveys were female with an average of 66% of respondents older than 51 years, 20% older than 41 years and 14% between 18-40 years of age, which is representative and consistent with 2018 Tasmanian Health Workforce data, which shows an aging nursing workforce (Table 1).

**Table 1:** Demographics

| Demographic variable | Classifications | Survey 1 |  | Survey 2 |  | Survey 3 |  |
| --- | --- | --- | --- | --- | --- | --- | --- |
|  |  | N | % | N | % | N | % |
| Age group | 18 - 30 | 46 | 6 | 41 | 7 | 29 | 7 |
|  | 31 - 50 | 194 | 28 | 140 | 26 | 118 | 27 |
|  | 51 + | 444 | 66 | 371 | 97 | 290 | 66 |
| Employee classification | Registered Nurse | 572 | 84 | 473 | 86 | 362 | 82 |
|  | Midwife | 40 | 6 | 26 | 5 | 26 | 6 |
|  | Enrolled Nurse | 56 | 8 | 32 | 6 | 38 | 9 |
|  | Assistant in Nursing | 8 | 1 | 8 | 1 | 5 | 1 |
|  | Not answered | 8 | 1 | 14 | 1 | 8 | 1 |
| Education Level | Undergraduate | 9 | 1.6 | 6 | 1 | 4 | 1 |
|  | Bachelor | 210 | 31 | 171 | 31 | 130 | 30 |
|  | Grad Certificate/ higher | 355 | 51 | 283 | 51 | 219 | 50 |
|  | Diploma /Hospital training certificate | 96 | 14 | 79 | 14 | 80 | 18 |
|  | Not answered | 14 | 1 | 14 | 1 | 6 | 1 |
| Region employed | Northern Tasmania | 125 | 18 | 112 | 20 | 97 | 22 |
|  | North West Tasmania | 42 | 6 | 14 | 2 | 63 | 14 |
|  | Southern Tasmania | 500 | 74 | 403 | 73 | 262 | 61 |
|  | Statewide | 8 | 1 | 9 | 1 | 11 | 1 |
|  | Not answered | 9 | 1 | 15 | 1 | 6 | 1 |
| Social situation | Living with family | 373 | 82 | 434 | 78 | 352 | 80 |
|  | Living with friends | 11 | 3 | 78 | 14 | 8 | 2 |
|  | Living alone | 65 | 14 | 78 | 14 | 69 | 16 |
|  | Other | 6 | 1 | 11 | 2 | 4 | 1 |
|  | Not answered | 229 | 33 | 15 | 1 | 6 | 1 |

Approximately 84% of the study sample were Registered Nurses (incorporating RN-Midwife), 17% Enrolled Nurses and 6% Midwives. The educational level of participants across all surveys shows 15% of participants hold a TAFE Diploma /Hospital training certificate, 31% of participants hold a bachelor’s degree and 50% a graduate certificate or higher. The state of Tasmania has 3 regions with 70% of study respondents living in the Southern region. Eighteen percent of respondents indicated they looked after COVID-19 positive patients at timepoint 1, dropping to 9% at timepoint 2 and 7% at timepoint 3. Living situations showed across all surveys, 80% of nurses live with family, 15% live alone, and 3% live with friends. The number of nurses who reported they were smokers remained consistent across all surveys, reporting a rate of 7%.

### Workplace predictors over time

Nurses and midwives were asked about a range of workplace issues specific to the COVID-19 pandemic. Fifty-five percent of respondents consistently felt ‘definitely confident’ or ‘moderately confident’ to provide competent care in the event they were deployed with only 9 percent of nurses reporting they did not feel confident across all timepoints. Sixty-nine percent of respondents felt adequately supported by their workplace team (colleagues and line manager) ‘all of the time’ or ‘most of the time’ across all timepoints with 27% indicating ‘some of the time’ or ‘never’. Fifty-four percent of respondents felt they were definitely providing quality patient care, 27% moderately and 10% only slightly or not at all. When asked if they have adequate information, clear communication, and guidelines about COVID-19, 72% of respondents reported ‘all of the time’ or ‘most of the time’ and only 25% indicating ‘some of the time’ or ‘never’. Respondents were asked if they were concerned about the access to personal protective equipment (PPE), 61% stated they were ‘very concerned’ or ‘moderately concerned’. Sixty-four percent of respondents indicated they had access to rapid COVID-19 testing ‘all the time’ or ‘most of the time’ across all timepoints. Concern around the access to PPE and rapid testing was slightly higher at timepoint.

### Psychological Outcomes

Table 2 shows mean scores for all respondents for surveys 1, 2 and 3. Scores show moderate to severe levels of anxiety and depression together with low mean scores of general stress/PTSD and insomnia across all timepoints. Levels of general stress / PTSD are highest at timepoint 1 then trend downward.

**Table 2:**
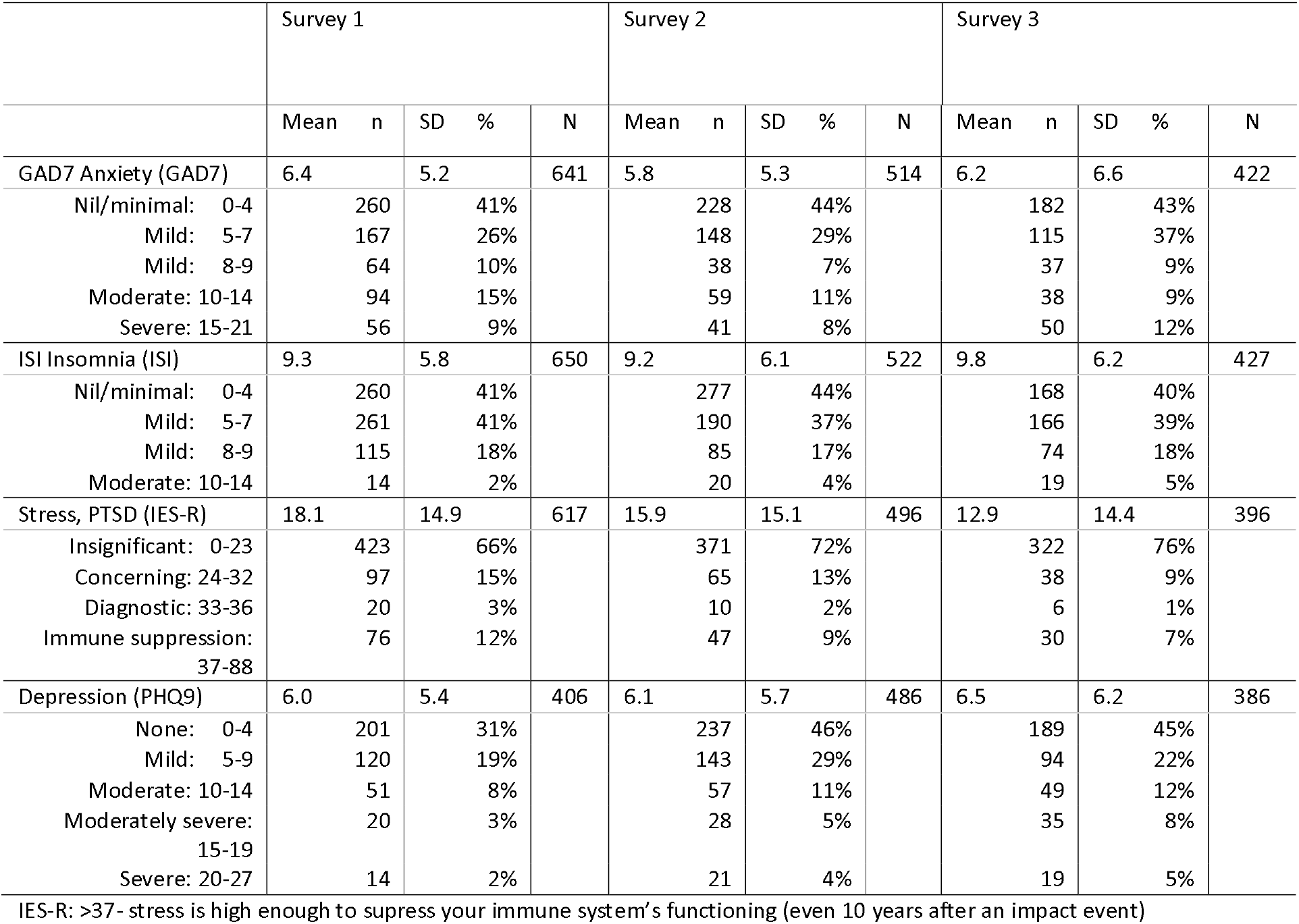
Psychological test mean (SD) scores, and numbers (%) in each classification range

While mean scores of general stress/PTSD scores indicate no clinical case, standard deviations indicate outliers or respondents with extremely high scores which well above the mean, indicating levels of general stress/PTSD of clinical concern. These outlier respondents will be described through the analysis the association of predictors and psychological outcomes.

### Trends in association of demographic variables and psychological outcomes

Table 3 shows significant negative relationships exist between level of education and insomnia throughout all study timepoints: when levels of education decrease, the levels of insomnia increase (clinically subthreshold levels).

**Table 3.** Trend of association between psychological scale and highest education level in the three surveys

| Scale | Age group | Survey | N | Mean | SD | OR for trend | 95%CI | P-value |
| --- | --- | --- | --- | --- | --- | --- | --- | --- |
| GAD7 | 18-29 | 1 | 33 | 9.5 | 6.4 | 0.62 | (0.46, 0.83) | 0.0015 |
|  | 30-49 |  | 182 | 7.1 | 5.3 |  |  |  |
|  | 50+ |  | 426 | 5.8 | 5.0 |  |  |  |
|  | 18-29 | 2 | 27 | 6.8 | 5.2 | 0.37 | (0.21, 0.63) | 0.0003 |
|  | 30-49 |  | 127 | 7.1 | 6.1 |  |  |  |
|  | 50+ |  | 360 | 5.3 | 4.8 |  |  |  |
|  | 18-29 | 3 | 20 | 8.4 | 6.3 | 0.56 | (0.30, 1.03) | 0.064 |
|  | 30-49 |  | 115 | 8.3 | 5.9 |  |  |  |
|  | 50+ |  | 287 | 5.2 | 5.2 |  |  |  |
| Scale | Education level | Survey | N | Mean | SD | OR for trend | 95%CI | P-value |
| Insom7 | Diploma/Certificate | 1 | 107 | 10.4 | 5.9 | 0.65 | (0.52, 0.92) | 0.0003 |
|  | Bachelor degree |  | 200 | 9.8 | 5.4 |  |  |  |
|  | Higher degree |  | 343 | 8.7 | 6.0 |  |  |  |
|  | Diploma/Certificate | 2 | 82 | 9.6 | 6.3 | 0.62 | (0.48, 0.80) | 0.0003 |
|  | Bachelor degree |  | 165 | 10.2 | 6.8 |  |  |  |
|  | Higher degree |  | 275 | 8.4 | 5.6 |  |  |  |
|  | Diploma/Certificate | 3 | 83 | 10.5 | 6.6 | 0.70 | (0.53, 0.92) | 0.011 |
|  | Bachelor degree |  | 126 | 10.1 | 5.9 |  |  |  |
|  | Higher degree |  | 218 | 9.3 | 6.2 |  |  |  |

Unadjusted mean (and standard deviation) scale value for insomnia subgroup; Odds ratio for trend denotes trend of adjusted association between psychological scale and education level (expressed as a z-score). Total score cut-off points: absence of insomnia (0–7), sub-threshold insomnia (8–14), moderate insomnia (15–21), and severe insomnia (22–28)

There were no significant relationships between employment classification, levels of education and outcomes of anxiety, depression, or general stress/PTSD.

There was significant relationships between age and anxiety; there were higher levels of anxiety among younger nurses across all timepoints (Table 3).

### Workplace COVID exposure

Study data examined associations between the psychological responses of nurses who worked on COVID-19 positive and negative wards. Data show only one significant relationship with anxiety - as the threat of COVID-19 increased; levels of anxiety increased to clinically moderate levels in Survey 1 only. There were no significant relationships with the other psychological outcome measures at any other timepoints.

### Relative impact of predictors on psychological outcomes

The relative impact of different predictors on the outcome scores (Tables 4 & 5) was estimated as odds ratios for trend using multivariate non-parametric regression.

**Table 4.** Relative impact of predictors on GAD7 and Insom7 scores in combined responses from all three surveys

| Predictor | OR for trend | 95%CI | P-value | Predictor | OR for trend | 95%CI | P-value |
| --- | --- | --- | --- | --- | --- | --- | --- |
| GAD7 |  |  |  | Insom7 |  |  |  |
| Current home/family stress | 2.25 | (1.88, 2.70) | <0.0001 | Current home/family stress | 1.75 | (1.47, 2.09) | <0.0001 |
| Poor clinical team support | 1.77 | (1.51, 2.09) | <0.0001 | Poor clinical team support | 1.51 | (1.28, 1.77) | <0.0001 |
| Female gender | 1.31 | (1.12, 1.53) | 0.0007 | Current financial stress | 1.37 | (1.13, 1.66) | 0.0015 |
| Future home/family stress | 1.25 | (1.05, 1.50) | 0.014 | Poor access to rapid tests | 1.27 | (1.09, 1.48) | 0.0026 |
| Future financial stress | 1.25 | (1.02, 1.52) | 0.028 | Concerned about PPE | 1.24 | (1.07, 1.44) | 0.0046 |
| Poor ability for quality care | 1.23 | (1.05, 1.43) | 0.0009 | Poor ability for quality care | 1.23 | (1.05, 1.44) | 0.0095 |
| Concerned about PPE | 1.23 | (1.05, 1.43) | 0.0009 | Future home/family stress | 1.22 | (1.02, 1.46) | 0.029 |
| Poor access to rapid tests | 1.12 | (0.96, 1.31) | 0.15 | Female gender | 1.12 | (0.96, 1.31) | 0.16 |
| Covid ward status | 1.10 | (0.95, 1.29) | 0.19 | Future financial stress | 1.10 | (0.90, 1.34) | 0.33 |
| Current financial stress | 1.10 | (0.95, 1.29) | 0.35 | Covid ward status | 1.08 | (0.94, 1.26) | 0.28 |
| Social situation | 0.99 | (0.84, 1.15) | 0.86 | Social situation | 1.04 | (0.89, 1.22) | 0.59 |
| Higher staff grade | 0.99 | (0.85, 1.16) | 0.94 | Intensity of exposure | 0.97 | (0.84, 1.13) | 0.71 |
| Public sector | 0.96 | (0.84, 1.09) | 0.52 | Smoking | 0.96 | (0.83, 1.11) | 0.58 |
| Intensity of exposure | 0.93 | (0.81, 1.08) | 0.37 | Higher staff grade | 0.94 | (0.81, 1.10) | 0.43 |
| Smoking | 0.89 | (0.77, 1.03) | 0.11 | Age | 0.89 | (0.75, 1.05) | 0.17 |
| Higher education level | 0.87 | (0.75, 1.02) | 0.098 | Adequacy of information | 0.83 | (0.71, 0.97) | 0.021 |
| Adequacy of information | 0.86 | (0.73, 1.00) | 0.055 | Public sector | 0.81 | (0.71, 0.92) | 0.0009 |
| Age | 0.56 | (0.48, 0.67) | <0.0001 | Higher education level | 0.67 | (0.57, 0.79) | <0.0001 |

Tables 4 and 5 show the relative impact of predictors on psychological outcomes and show there are strong relationships consistently across many psychological outcomes. Describing effects over time will help contextualise the responses and enable some insight into how nurses and midwives adapt to the pandemic.

**Table 5.** Relative impact of predictors on IESR and PHQ scores in combined responses from all three surveys

| Predictor | OR for trend | 95%CI | P-value | Predictor | OR for trend | 95%CI | P-value |
| --- | --- | --- | --- | --- | --- | --- | --- |
| IESR22 |  |  |  | PHQ9 |  |  |  |
| Current home/family stress | 2.27 | (1.91, 2.71) | <0.0001 | Current home/family stress | 1.78 | (1.44, 2.20) | <0.0001 |
| Concerned about PPE | 1.52 | (1.31, 1.76) | <0.0001 | Poor clinical team support | 1.59 | (1.32, 1.92) | <0.0001 |
| Poor clinical team support | 1.47 | (1.27, 1.70) | <0.0001 | Future home/family stress | 1.35 | (1.09, 1.68) | 0.0059 |
| Female gender | 1.37 | (1.19, 1.59) | <0.0001 | Female gender | 1.30 | (1.08, 1.57) | 0.0064 |
| Future home/family stress | 1.32 | (1.12, 1.52) | 0.0013 | Current financial stress | 1.29 | (1.02, 1.62) | 0.033 |
| Future financial stress | 1.30 | (1.08, 1.56) | 0.0059 | Social situation | 1.27 | (1.06, 1.52) | 0.010 |
| Current financial stress | 1.25 | (1.04, 1.49) | 0.015 | Concerned about PPE | 1.26 | (1.05, 1.51) | 0.012 |
| Poor ability for quality care | 1.22 | (1.06, 1.41) | 0.0066 | Poor ability for quality care | 1.18 | (0.98, 1.42) | 0.075 |
| Covid ward status | 1.20 | (1.04, 1.38) | 0.013 | Future financial stress | 1.16 | (0.91, 1.46) | 0.22 |
| Public sector | 1.15 | (1.02, 1.29) | 0.026 | Covid ward status | 1.11 | (0.93, 1.34) | 0.24 |
| Social situation | 1.12 | (0.97, 1.30) | 0.11 | Poor access to rapid tests | 1.06 | (0.88, 1.28) | 0.52 |
| Intensity of exposure | 1.00 | (0.87, 1.15) | 0.97 | Intensity of exposure | 1.06 | (0.89, 1.26) | 0.51 |
| Age | 0.95 | (0.82, 1.10) | 0.50 | Lower staff grade | 0.97 | (0.81, 1.16) | 0.72 |
| Poor access to rapid tests | 0.92 | (0.80, 1.07) | 0.28 | Higher education level | 0.92 | (0.76, 1.11) | 0.39 |
| Smoking | 0.87 | (0.76, 0.99) | 0.039 | Smoking | 0.88 | (0.77, 1.10) | 0.34 |
| Lower staff grade | 0.86 | (0.75, 0.99) | 0.041 | Public sector | 0.88 | (0.71, 1.09) | 0.25 |
| Higher education level | 0.84 | (0.72, 0.97) | 0.016 | Adequacy of information | 0.75 | (0.62, 0.92) | 0.0045 |
| Adequacy of information | 0.76 | (0.66, 0.89) | 0.0004 | Age | 0.61 | (0.50, 0.75) | <0.0001 |

### Workplace predictors and psychological outcomes

#### 1. Effect of ‘CONCERN ABOUT ACCESS TO PPE’ across all psychological domains

**Figure 1.**
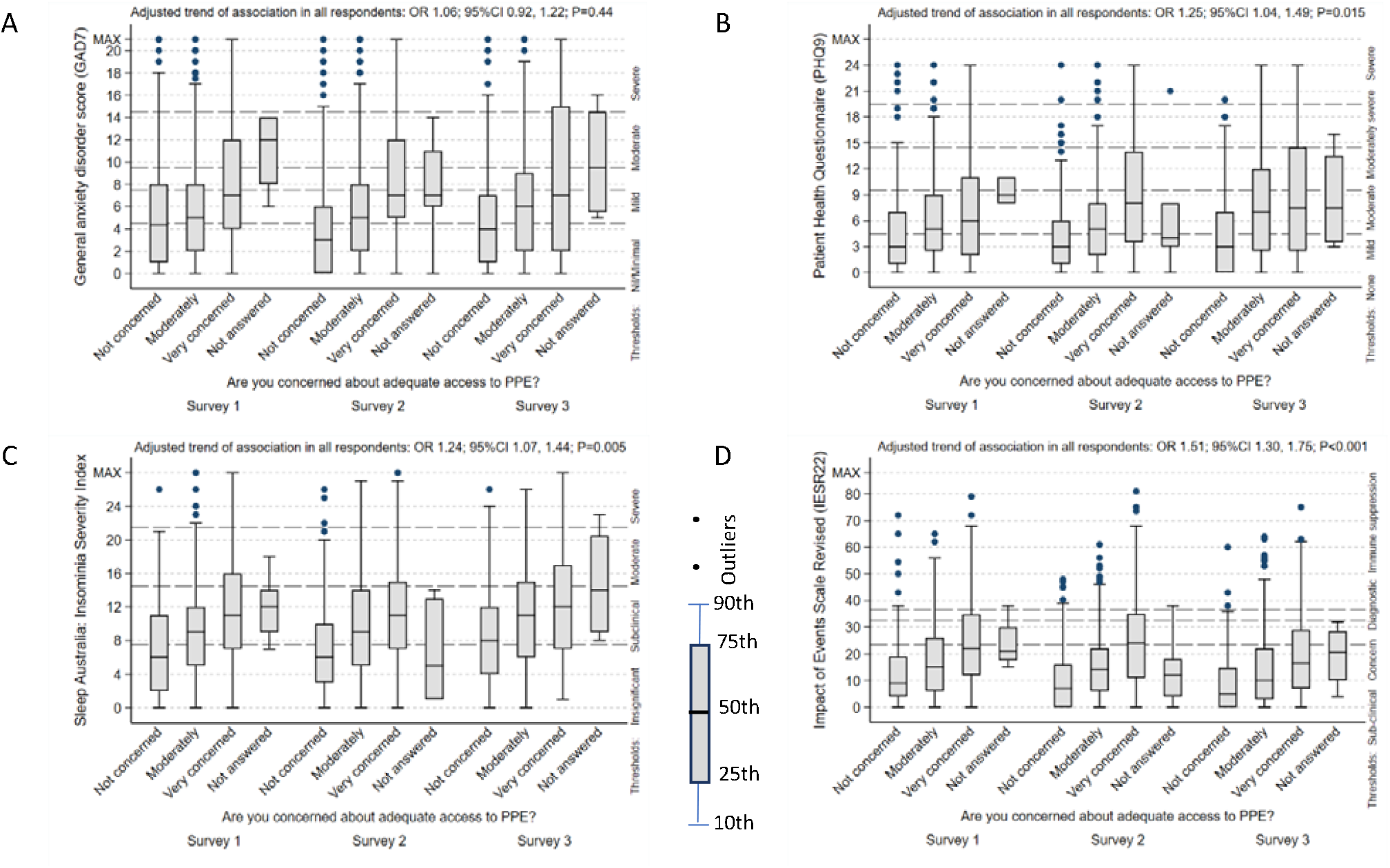
Effect of concern about access to PPE across all psychological domains. Note that the boxplots are nor adjusted for the other covariates, whilst the odds ratios for trend are adjusted for other covariates.

Box plots 1A, 1B, 1C and 1D show the strong and consistent statistical relationship between the effect of concern about adequate access to PPE across all psychological domains, showing as concern about adequate access to PPE increase, so do the levels of anxiety, depression, insomnia (all reach clinically severe levels) and general stress/PTSD which reaches such severe levels which can compromise immune system function. Though the relationship with anxiety does not show significance when the 3 surveys are taken together, the trends in Survey 2 at 3 months (odds ratio for trend 1.43; 95%CI 1.12, 1.82; P=0.004) and Survey 3 at 12 months (OR for trend 1.20; 95%CI 0.93, 1.56; P=0.17) compared to Survey 1 at the start of the pandemic (OR for trend 1.05; 95%CI 0.85, 1.31; P=0.64) are consistent with the other psychological outcome measures, but insignificant initially reaching a maximum at 3 months and then diminishing. Mean scores of insomnia relating to PPE show an upwards trend up over time, reaching highest levels at 12 months (OR for trend 1.43; 95%CI 1.08, 1.90; P=0.013).

#### 2. Effect of CONCERN FOR LACK OF CLNICAL TEAM SUPPORT across all psychological domains

**Figure 2.**
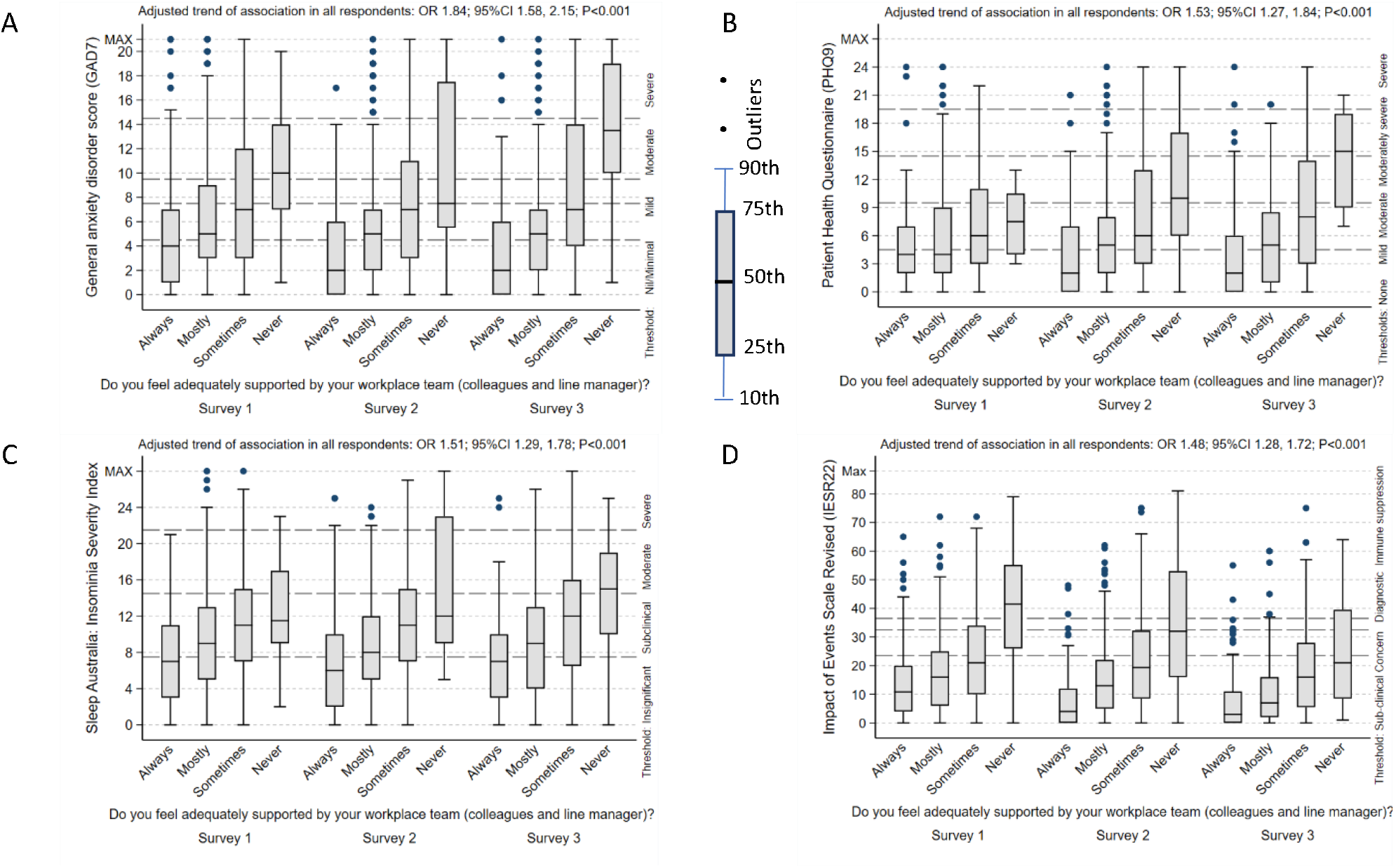
Effect of concern for lack of clinical team support across all psychological domains. Note that the boxplots are nor adjusted for the other covariates, whilst the odds ratios for trend are adjusted for other covariates.

Box plots 2A, 2B, 2C and 2D show the strong and consistent statistical relationship between the effect of concern of adequate support from the workplace team across all psychological domains, showing as concern for adequate support from the workplace team increase, so do the levels of anxiety, depression, insomnia (all reach clinically severe levels) and general stress/PTSD which reach such severe levels which can compromise immune system function. Of note are the levels of anxiety and depression which increase over time: GAD7 (initial OR for trend 1.69; 95%CI 1.35, 2.12; P<0.0001), (3month OR for trend 1.65; 95%CI 1.28, 2.13; P=0.0001), (12 month OR for trend 2.28; 95%CI 1.74, 2.98; P<0.0001); ISI-7 (initial OR for trend 1.26; 95%CI 1.00, 1.60; P0.054), (3month OR for trend 1.70; 95%CI 1.30, 2.23; P=0.0001), (12 month OR for trend 2.28; 95%CI 1.74, 2’98; P<0.0001); IESR-22 (initial OR for trend 1.69; 95%CI 1.35, 2.12; P<0.0001), (3month OR for trend 1.65; 95%CI 1.28, 2.13; P=0.0001), (12 month OR for trend 1.86; 95%CI 1.39, 2.48; P<0.0001).

#### 3. Effect of ‘CONCERN FOR ADEQUATE INFORMATION’ across anxiety, depression and general stress/PTSD psychological domains

**Figure 3.**
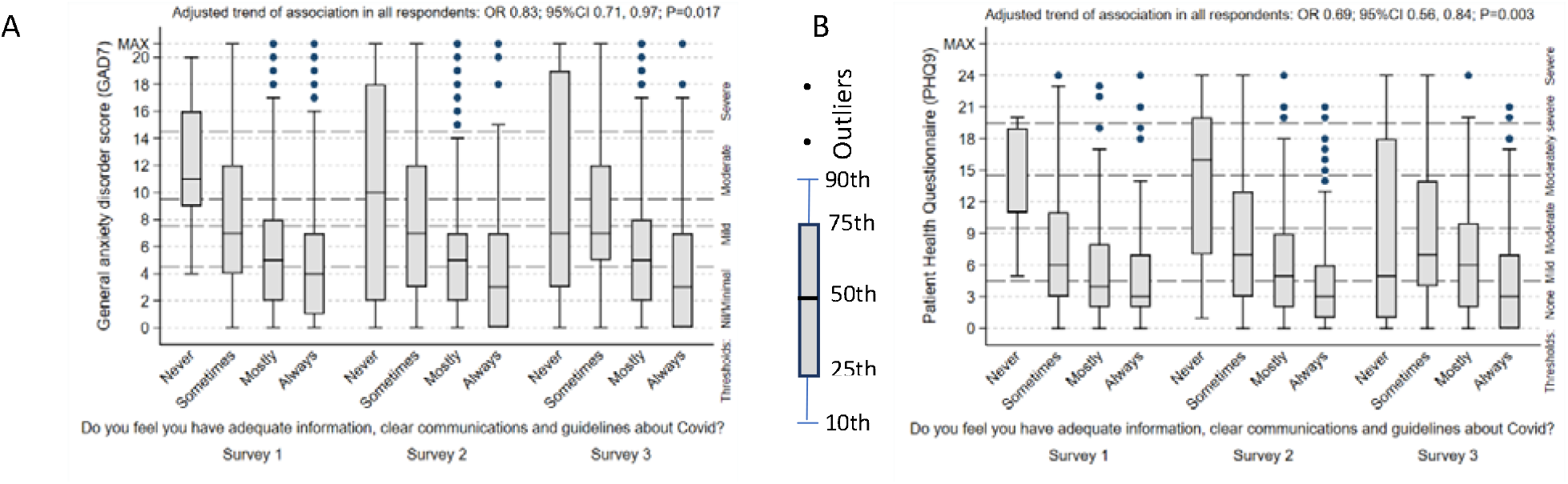
Effect of concern for adequate information across all psychological domains. Note that the boxplots are nor adjusted for the other covariates, whilst the odds ratios for trend are adjusted for other covariates.

Box plots 3A, and 3B, show the negative statistical relationship between adequacy of information and anxiety and depression domains, showing as concern about adequate information increases, so do the levels of anxiety and depression. This effect diminishes over time, more for GAD7 scores than for PHQ9 scores: GAD7 (initial OR for trend 0.75; 95%CI 0.60, 0.93; P=0.0089), (3month OR for trend 0.88; 95%CI 0.68, 1.12; P=0.31), (12 month OR for trend 0.92; 95%CI 0.70, 1.21; P=0.56); PHQ9 (initial OR for trend 0.60; 95%CI 0.44, 0.83; P0.0019), (3month OR for trend 0.75; 95%CI 0.56, 1.01; P=0.059), (12 month OR for trend 0.71; 95%CI 0.51, 1.00; P=0.051);.

A strong focal relationship exists between concern for adequate information and general stress/PTSD at the 3-month survey (Table 6); as levels of concern increase, so do levels of general stress/PTSD, with mean scores reaching levels of clinical concern.

**Table 6.** Trend of association between IESR scale and lack of adequate information in 3 surveys.

| Scale | Adequacy of information | Survey | N | Mean<sup>2</sup> | SD | OR for trend | 95%CI | P-value |
| --- | --- | --- | --- | --- | --- | --- | --- | --- |
| IESR22 | All of the time | 1 | 13 | 28.4 | 11.1 | 0.80 | (0.64, 1.00) | 0.051 |
|  | Most of the time |  | 171 | 23.8 | 16.9 |  |  |  |
|  | Some of the time |  | 270 | 15.5 | 13.1 |  |  |  |
|  | None of the time |  | 156 | 14.6 | 13.5 |  |  |  |
|  | All of the time | 2 | 8 | 34.1 | 24.2 | 0.62 | (0.48, 0.81) | 0.0004 |
|  | Most of the time |  | 101 | 22.0 | 17.5 |  |  |  |
|  | Some of the time |  | 238 | 15.6 | 13.9 |  |  |  |
|  | None of the time |  | 146 | 10.2 | 12.2 |  |  |  |
|  | All of the time | 3 | 11 | 28.5 | 25.2 | 0.87 | (0.65, 1.16) | 0.35 |
|  | Most of the time |  | 83 | 16.5 | 16.5 |  |  |  |
|  | Some of the time |  | 183 | 13.0 | 13.5 |  |  |  |
|  | None of the time |  | 116 | 8.6 | 10.7 |  |  |  |

IES-R severity: 24-33- PTSD is of clinical concern; 33-37- Probable PTSD diagnosis; >37- stress is high enough to suppress your immune system's functioning (even 10 years after an impact event); OR – Odds ratio for trend estimated by ordered logistic regression adjusted for covariates.

#### 4. Effect of ‘CONCERN FOR PROVIDING QUALITY CARE’ across anxiety, depression domains

A strong focal relationship is seen at 3 months in the area of concern for providing quality care and anxiety and depression. As the concern for the provision of quality care increases so does the levels of anxiety and depression, both reaching sever levels (Table 7).

**Table 7.** Trend of association between GAD and PHQ scale and poor-quality care delivery in the 3 surveys

| Scale | Ability for quality care | Survey | N | Mean<sup>2</sup> | SD | OR for trend | 95%CI | P-value |
| --- | --- | --- | --- | --- | --- | --- | --- | --- |
| GAD7 | Definitely | 1 | 340 | 11.0 | 7.3 | 1.14 | (0.92, 1.15) | 0.23 |
|  | Moderately |  | 181 | 12.6 | 7.2 |  |  |  |
|  | Slightly |  | 54 | 12.7 | 7.2 |  |  |  |
|  | Not at all |  | 14 | 14.5 | 5.6 |  |  |  |
|  | Definitely | 2 | 257 | 14.2 | 5.5 | 1.38 | (1.08, 1.77) | 0.011 |
|  | Moderately |  | 164 | 16.6 | 6.0 |  |  |  |
|  | Slightly |  | 48 | 19.5 | 8.0 |  |  |  |
|  | Not at all |  | 9 | 24.4 | 8.7 |  |  |  |
|  | Definitely | 3 | 221 | 14.3 | 6.2 | 1.24 | (0.94, 1.62) | 0.12 |
|  | Moderately |  | 127 | 17.7 | 6.5 |  |  |  |
|  | Slightly |  | 38 | 18.1 | 6.1 |  |  |  |
|  | Not at all |  | 10 | 22.4 | 8.6 |  |  |  |
| PHQ9 | Definitely | 1 | 226 | 14.8 | 5.9 | 1.08 | (0.80, 1.47) | 0.59 |
|  | Moderately |  | 120 | 16.6 | 5.8 |  |  |  |
|  | Slightly |  | 28 | 17.4 | 7.1 |  |  |  |
|  | Not at all |  | 10 | 17.0 | 5.4 |  |  |  |
|  | Definitely | 2 | 241 | 14.2 | 5.2 | 1.40 | (1.03, 1.89) | 0.028 |
|  | Moderately |  | 157 | 16.8 | 6.6 |  |  |  |
|  | Slightly |  | 47 | 19.9 | 8.0 |  |  |  |
|  | Not at all |  | 7 | 22.9 | 6.7 |  |  |  |
|  | Definitely | 3 | 207 | 14.7 | 6.2 | 1.02 | (0.73, 1.43) | 0.88 |
|  | Moderately |  | 112 | 18.6 | 7.5 |  |  |  |
|  | Slightly |  | 35 | 18.8 | 6.6 |  |  |  |
|  | None of the time |  | 7 | 22.9 | 6.9 |  |  |  |

PHQ severity: 0-4 none, 5-9 mild, 10-14 moderate, 15-19 moderately severe, 20-27 severe. GAD severity: 0-4 none to minimal, 5-7 mild (recommended to monitor symptoms), 8-9 mild though likely to be diagnosed with an anxiety disorder, 10-14 moderate symptoms are clinically significant, 15-21 severe symptoms warranting active treatment; OR – Odds ratio for trend estimated by ordered logistic regression adjusted for covariates.

There were no significant relationships identified for both predictors, ‘access to rapid COVID-19 testing’ and ‘confidence to provide competent care in the event they were deployed’ across all psychological domains.

### Social predictors and psychological outcomes

#### 5. Effect of ‘CURRENT (and FUTURE) HOME AND FAMILY STRESS’ across all psychological domains

Nurses were asked to what degree does COVID-19 affect current (and future) home and family stress. Box plots 5A, 5B, 5C and 5D show the strong and consistent statistical relationship between the effect of current home and family stress across all psychological domains, showing as current home and family stress increase, so do the levels of anxiety, depression, insomnia (clinically severe levels) and general stress/PTSD which reach such severe levels which can compromise immune system function. The effect for anxiety and depression continues increases over time with highest levels of anxiety and depression occurring at timepoint 3.

There were no significant relationships across any psychological domains associated with future home and family stress.

**Figure 4.**
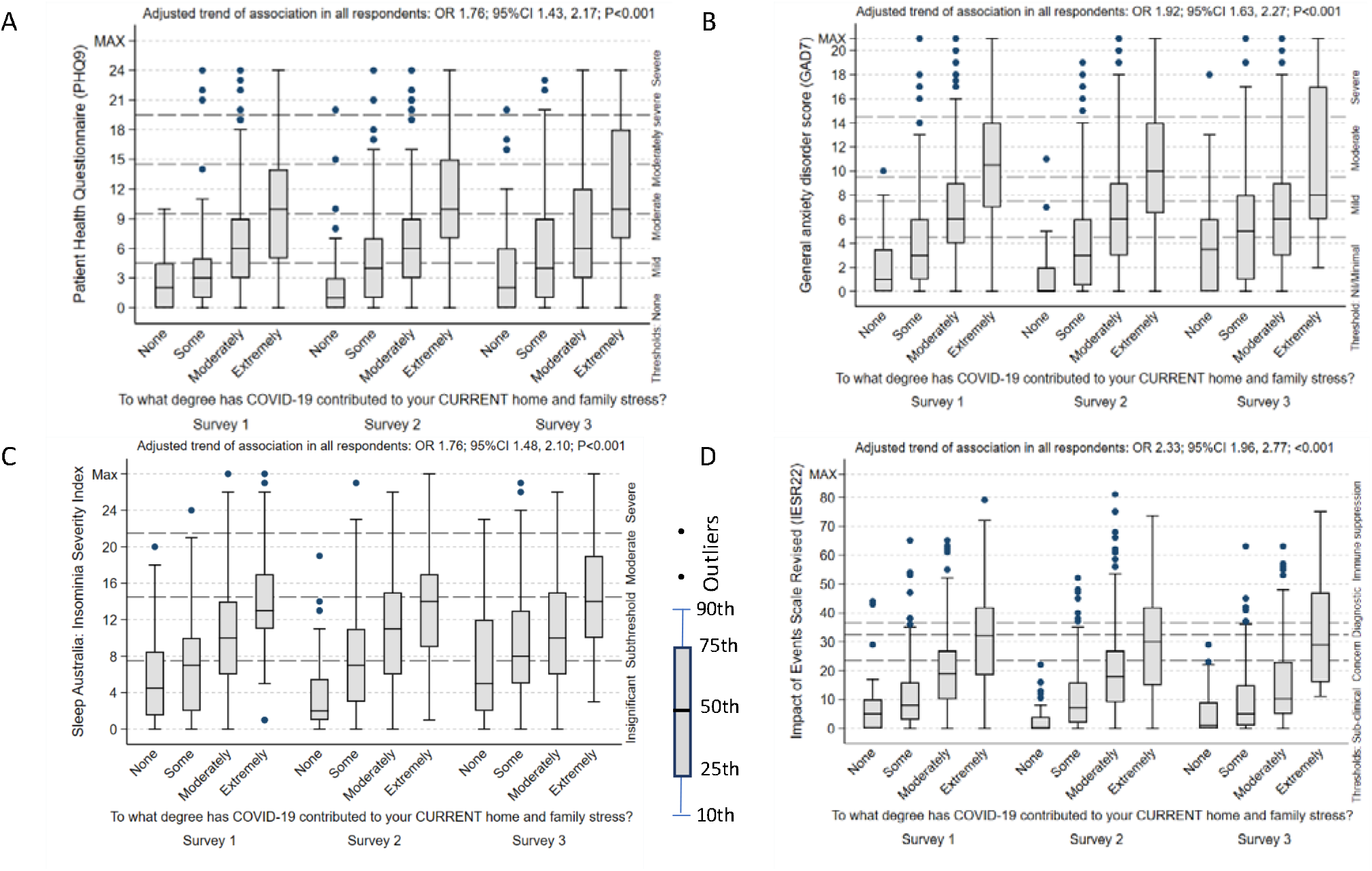
Effect of current home and family stress across all psychological domains. Note that the boxplots are nor adjusted for the other covariates, whilst the odds ratios for trend are adjusted for other covariates.

#### 6. Effect of CURRENT (AND FUTURE) FINANCIAL STRESS’ across all psychological domains

**Figure 5.**
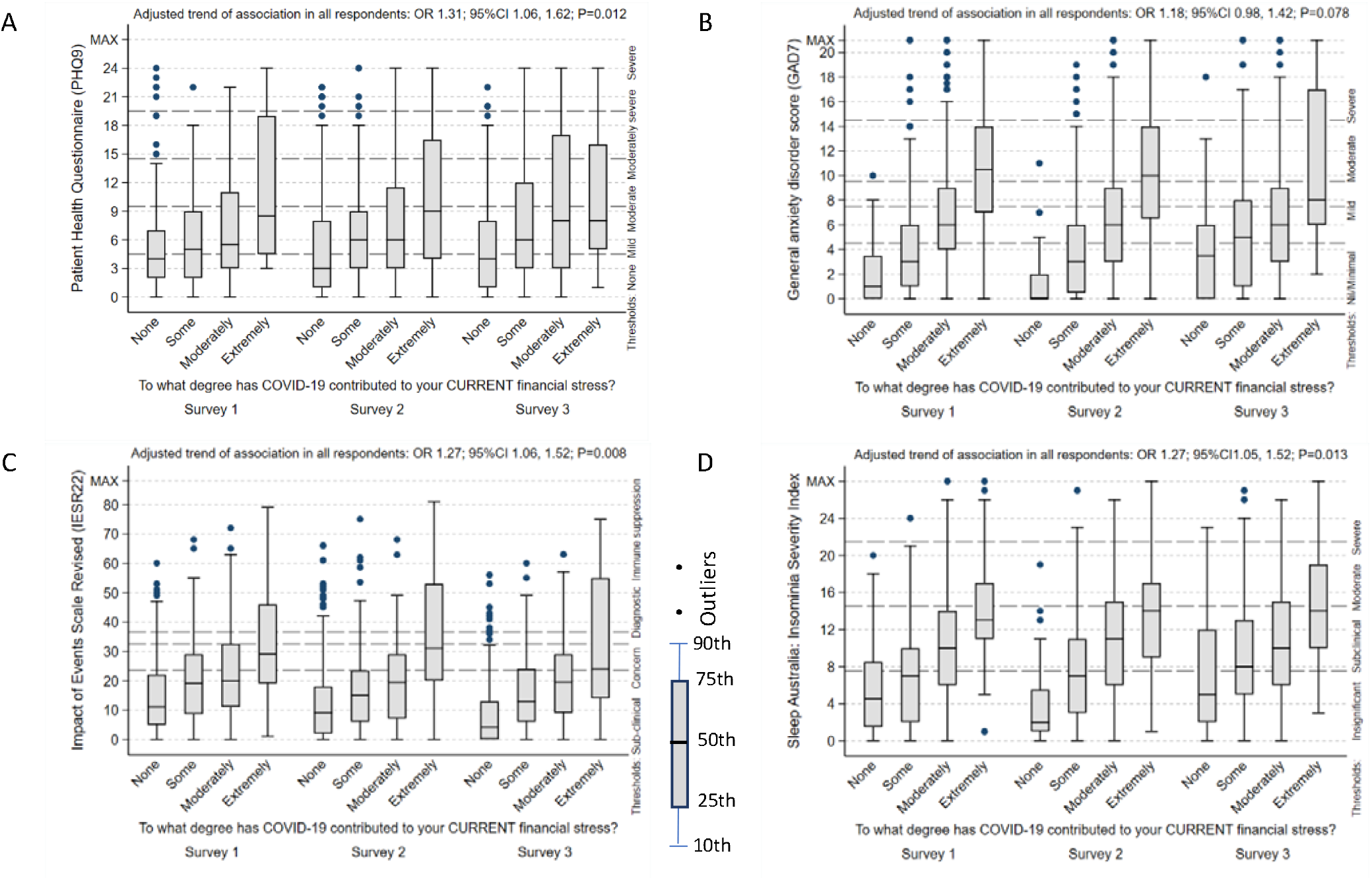
Effect of current (and future) financial stress’ across anxiety, depression and general stress/PTSD domains Note that the boxplots are nor adjusted for the other covariates, whilst the odds ratios for trend are adjusted for other covariates.

Nurses were asked to what degree does COVID-19 affect current (and future) financial stress. Box plots 6A, 6B and 6C show the strong and consistent statistical relationship between the effect of current financial stress across anxiety, depression and general stress/PTSD psychological domains, showing as financial stress increase, so do the levels of anxiety, depression, and general stress/PTSD which reach such severe levels which can compromise immune system function. Results show (Table 8) increasing statistical relationships and high scores for general stress/PTSD, particularly at 3 months and 12 months. Strong associations together with high mean scores are apparent for depression (3 months and 12 months) anxiety (12 months) and insomnia at 12 months. Trend patterns throughout all the graphs are similar, showing increasing median scores as the study progresses. Future financial stress was associated with anxiety (OR for trend 1.64; 95%CI 1.21, 2.23; P=0.0015), and general stress/PTSD (OR for trend 1.58; 95%CI 1.18, 2.11; P=0.0022) psychological domains at the initial survey, but no associations were found with any of the psychological domains at 3 months or 12 months.

**Table 8.** Trend of association between psychological outcomes and scale and CURRENT FINANCIAL STRESS in the three surveys

| Scale | Current financial stress | Survey | N | Mean<sup>2</sup> | SD | OR for trend | 95%CI | P-value |
| --- | --- | --- | --- | --- | --- | --- | --- | --- |
| GAD7 | Not at all | 1 | 362 | 10.6 | 6.8 | 0.97 | (0.73, 1.29) | 0.82 |
|  | Some |  | 142 | 11.9 | 7.3 |  |  |  |
|  | Moderately |  | 100 | 13.6 | 7.8 |  |  |  |
|  | Extremely |  | 37 | 14.3 | 8.4 |  |  |  |
|  | Not at all | 2 | 292 | 14.6 | 5.9 | 1.12 | (0.82, 1.54) | 0.47 |
|  | Some |  | 133 | 16.8 | 5.9 |  |  |  |
|  | Moderately |  | 63 | 17.0 | 6.2 |  |  |  |
|  | Extremely |  | 26 | 21.3 | 8.8 |  |  |  |
|  | Not at all | 3 | 265 | 14.2 | 5.7 | 1.59 | (1.02, 2.39) | 0.042 |
|  | Some |  | 79 | 17.5 | 6.4 |  |  |  |
|  | Moderately |  | 58 | 18.9 | 7.6 |  |  |  |
|  | Extremely |  | 20 | 21.7 | 7.2 |  |  |  |
| Insom7 | Not at all | 1 | 368 | 8.2 | 5.6 | 1.27 | (0.95, 1.69) | 0.11 |
|  | Some |  | 143 | 9.9 | 5.7 |  |  |  |
|  | Moderately |  | 102 | 11.1 | 5.6 |  |  |  |
|  | Extremely |  | 37 | 13.6 | 6.3 |  |  |  |
|  | Not at all | 2 | 296 | 7.9 | 5.8 | 1.18 | (0.86, 1.62) | 0.30 |
|  | Some |  | 137 | 10.0 | 5.7 |  |  |  |
|  | Moderately |  | 63 | 11.2 | 6.2 |  |  |  |
|  | Extremely |  | 26 | 14.2 | 6.8 |  |  |  |
|  | Not at all | 3 | 269 | 8.4 | 5.8 | 2.12 | (1.39, 3.24) | 0.0005 |
|  | Some |  | 80 | 11.2 | 5.8 |  |  |  |
|  | Moderately |  | 58 | 12.2 | 6.2 |  |  |  |
|  | Extremely |  | 20 | 16.0 | 6.2 |  |  |  |
| IESR22 | Not at all | 1 | 348 | 14.2 | 12.6 | 1.07 | (82, 1.41) | 0.61 |
|  | Some |  | 136 | 20.4 | 14.8 |  |  |  |
|  | Moderately |  | 96 | 22.6 | 15.7 |  |  |  |
|  | Extremely |  | 37 | 32.1 | 19.1 |  |  |  |
|  | Not at all | 2 | 284 | 12.5 | 12.9 | 1.38 | (1.01, 1.88) | 0.044 |
|  | Some |  | 129 | 17.1 | 14.6 |  |  |  |
|  | Moderately |  | 58 | 19.5 | 14.8 |  |  |  |
|  | Extremely |  | 25 | 34.4 | 23.8 |  |  |  |
|  | Not at all | 3 | 248 | 8.4 | 10.6 | 1.97 | (0.31, 2.96) | 0.0011 |
|  | Some |  | 73 | 16.4 | 13.9 |  |  |  |
|  | Moderately |  | 56 | 21.4 | 15.5 |  |  |  |
|  | Extremely |  | 19 | 31.6 | 23.6 |  |  |  |
| PHQ9 | Not at all | 1 | 238 | 14.5 | 5.4 | 1.34 | (0.91, 1.98) | 0.14 |
|  | Some |  | 83 | 15.8 | 5.4 |  |  |  |
|  | Moderately |  | 63 | 17.7 | 6.8 |  |  |  |
|  | Extremely |  | 22 | 21.0 | 8.8 |  |  |  |
|  | Not at all | 2 | 279 | 14.8 | 6.0 | 1.17 | (0.82, 1.67) | 0.38 |
|  | Some |  | 127 | 16.6 | 6.0 |  |  |  |
|  | Moderately |  | 56 | 17.4 | 6.9 |  |  |  |
|  | Extremely |  | 24 | 20.9 | 8.3 |  |  |  |
|  | Not at all | 3 | 241 | 14.6 | 5.9 | 1.67 | (1.04, 2.66) | 0.033 |
|  | Some |  | 71 | 17.8 | 6.7 |  |  |  |
|  | Moderately |  | 55 | 19.4 | 8.4 |  |  |  |
|  | Extremely |  | 19 | 21.4 | 8.4 |  |  |  |

PHQ severity: 0-4 none, 5-9 mild, 10-14 moderate, 15-19 moderately severe, 20-27 severe. GAD severity: 0-4 none to minimal, 5-7 mild (recommended to monitor symptoms), 8-9 mild though likely to be diagnosed with an anxiety disorder, 10-14 moderate symptoms are clinically significant, 15-21 severe symptoms warranting active treatment. IES-R severity: 24-33- PTSD is of clinical concern; 33-37- Probable PTSD diagnosis; >37- stress is high enough to suppress your immune system's functioning (even 10 years after an impact event). ISI severity: absence of insomnia (0-7), sub-threshold insomnia (8-14), moderate insomnia (15-21), and severe insomnia (22-28)

### Thematic Analysis

A thematic approach to analysing narratives was used to look for patterns, meaning and potential points of interest in the data and ensures an honest interpretation is maintained throughout the by constant reference to the raw data source(48). Nurses and midwives discussed their perceptions, experiences and memories of their COVID-19 experience at each time point. Narratives are subjective and may be influenced and altered with time. Narratives contained both factual and emotional memories, some explicit often chronologically pieced together.

Narrations were broadly separated into 5 themes:

1. Communication, advice and planning,
2. Vulnerability and accessibility of care,
3. Lack of social support, isolation and disconnect,
4. Work-related pressures
5. Positivity and reassurance.

#### Theme 1. COMMUNICATION, ADVICE AND PLANNING

Narratives uncovered nurses’ frustration around communication, advice, and planning. They included lack of clear communication, inconsistent advice, information, and planning with staff forced to change work practices in an attempt to reduce density of staff and patients in clinical areas. There was confusion around the use and meaning of evolving COVID-19 terminology, the rapidly changing directions and inconsistent enforcement of infection control procedures including access to supplies and training and use of PPE. There was frustration and confusion related to changes to hospital infrastructure and associated models of care, staff leave entitlements such as pandemic leave and the perceived lack of organisational support, all of which were changing almost daily.

*“Our daily lives are currently saturated with Covid19 from all sources, there is very little in the way of relief. We are expected to know so much, but it’s a rapidly moving feast. ID T1200”*

*“It was farcical – to keep 1*.*5 meters away from patients and staff in clinical practice was impossible. The process of patient handover became disjoint and dangerous. Patients were isolated until negative screening swabs came back which meant constantly moving patients around the ward and terminal cleaning bedspaces to accommodate this. “ ID T264*

With the lack of clear communication with staff, many nurses turned to alternative sources of information such as the general and social media. The initial months in Tasmania has been described by respondents as ‘feeling unprepared’ and ‘disjoint with a lack of organised health planning’, causing nurses to feel ‘confused, frightened, and unsupported’. There was lack of advice around what was considered appropriate regarding work and the risk to family, and the potential harm they posed as a viral vector. Professional and personal communications were blurred leaving staff feeling unsupported and perpetuated the spread of inaccurate information.

*“It was hard to get or discuss any clear information each day. The [Nurse Unit Managers] NUMs stopped having ward meetings and started writing newsletters once a month. The rest we read in the [local newspaper]. We knew things were changing constantly but had no idea what we should be doing. We all just did whatever we thought was a reasonable thing*.*” ID T1312*

There was much role change with new administrative or managerial positions created as part of the COVID-19 taskforce. Adopting new roles during an emergency, developing new lines of authority and accountability without adequate training provided many unique challenges.

*“The greatest stress for me is feeling unsupported by management and their refusal to acknowledge their appalling lack of communication. Being almost yelled at down the phone and told emails have been sent advised of change of my role at work. [I am] upset that I am expected to put my clients on the back burner to support others*.*” ID T1238*

*“I feel poorly supported by nursing management…*.. *I feel they are simply over-promoted nurses who lack management skills. They fail to advocate for us and for patients or deal with bullying and other unprofessional behaviours. This has contributed enormously to my anxiety and led me to requiring medical assistance*.*” ID T1243*

Many highlighted problems managing their existing workload, expressing feelings of failure and doubt about their capability to take on more while others expressed guilt if unable to work. There were grief reactions in response to the chaotic losses taking place. Nurses and midwives worried the current system did not have the surge capability or capacity to successfully manage the COVID-19 pandemic.

*“The current stress of nurses, staff and colleagues within the acute care facilities at this time does not stem from the COVID pandemic directly, but indirectly. The stress and workload demand that stems from the influx of highly acute patients and high level of patient flow and bed block is now daily life or “normal”. If there was an outbreak in Tasmania of COVID where would all the current ICU patients and respiratory patients go, if all of a sudden it was needed to support/ treat COVID patients? “ ID T1412*

*“I worry (and still do sometimes) for the staff and their wellbeing, more because I feel it would have been terrible for those involved. My initial reactions were relief because I was removed from the outbreak, and later guilt because of that relief, but also because I wasn’t able to lend a hand when people were working hours on end or make it better for those involved*.*” ID T1273*

#### Theme 2. VULNERABILITY AND ACCESSIBILITY OF CARE

Nurses felt ill-prepared and vulnerable as frontline health care workers. Perceived vulnerabilities stemmed from inadequate training, inadequate staff support protocols, insufficient PPE supplies, perceived lack of faith in the organisations’ infection control procedures, comorbid physical health conditions and older age. The organisation attempted to meet a need for consistent PPE training by providing a PPE training video which became both a support and a stressor. It was supportive as it was available on demand, however it also became a stressor for many clinicians, where PPE use was unfamiliar territory.

*“… There was no PPE for training. We don’t use PPE day-to-day so it was all new. We were told to watch a video*.*” ID T2374*

*“My ward has no changing facilities or showering facilities and minimal staff facilities. All staff are expected to change in a toilet cubicle (we have 1 toilet) and there’s only a sink in the cubicle where they can clean themselves after the shift. I’m informed that senior staff have discussed with the ward manager the lack of facilities and they were basically told it is what it is*.*” ID T3384*

Just like the general population, nurse’s personal lifestyles were impacted by the pandemic with gym closures and limited social opportunities which impacted on their ability to cope with the ever-changing and threatening environment. Negative coping mechanisms such as substance misuse increased alcohol and over-eating were evident creating psychological distress and precipitated mental health disorders.

*“This pandemic has taken away my coping mechanisms. I am finding it difficult to create new ones when all the rules are so strict*.*” ID T2610*

*“In the last two weeks have had a depressive episode related to my mental health illness not related to the COVID pandemic but the COVID pandemic has added another stressor to the mix*.*” ID: T251*

*“A glass or two of wine have become a daily in my house just to take the edge off*.*” ID T2473*

Nurses were conflicted with managing the welfare of their family unit with the welfare of their patients. Nurses with physical comorbidities or were immunocompromised, deemed high-risk, and those with young children or elderly family felt vulnerable, with many immediately withdrawing from work to reduce the risk of potential transmission.

*“I have immunosuppressed people in my immediate family and my partner works with an extremely vulnerable demographic so this has created stress by not being able to see my family as well as feeling a heightened sense of responsibility and pressure to ensure I do not get COVID to pass onto my partner who could then pass it on to their mega vulnerable patients. I am also petrified of the media and how they would villainise me and/or my partner if I would become positive as they have done to my NW colleagues*.*” ID: T2188*

Many older nurses either came to work, feeling scared, vulnerable, having to take on extremely busy workloads or withdrew from the workforce, precipitating many retire. This caused a sudden loss of experienced nurses from the workforce.

*“…*.. *[I am] concerned about the level of risk we are put under every day at work. What if I catch a superbug, get abused, night shifts and body clock/diet dysfunction, other influenza-like illnesses while we only seem to concentrate on COVID19. It been a hard few months*.*” ID T482*

*“I think that this approach of ‘oh we’ll send out a brochure or tell people about the EAP [Employee Assistance Program] or what have you is totally missing the point. Healthcare workers need practical assistance with the difficulties caused by this crisis - childcare, support to take breaks and leave on time, release of pressure to work extra shifts, or hazard pay to provide adequate compensation for the added risk and stress. And literally just a genuine acknowledgement from leadership that this is hard, that it is having flow on effects to our families, and that they’re grateful. And tacking that sentence onto the end of an email is NOT satisfactory*.*” ID T381*

Maintenance of their own personal health proved difficult. Nurses also endured the difficulties accessing medical and health care treatment, moving to telehealth for their own healthcare. There were many comments of fear and stress as nurses waited for COVID-19 test results; fearful of media portrayal as a COVID-19 positive health care worker. Nurses were aware their workplace decisions could potentially result in front-page news headlines if there was nosocomial spread. The information or directives which they based these clinical decisions upon did not exist.

To society, nurses were both heroes and villains. Nurses endured societal stigma, regarded as COVID-19 vectors and felt vulnerable travelling to and from work in uniform enduring public verbal attacks.

*“I am annoyed that today I am regarded as a hero yet I am just doing the same job I did yesterday” IDT171*

*“I am afraid to use public transport and walk for exercise alone for fear of attacks*.*” ID T323*

*“…*.*a lot of my mental health issues are less related to covid and more to do with personal circumstances, raising young children, social isolation, lack of external support, self-esteem issues*.*” ID T2153*

*“I feel there is a current hunt from the media in trying to establish blame*.*” ID T1424*

The vulnerability of patients caused much stress and anxiety for nurses. Nurses were sensitive to the disruption of patient treatments and services and the difficulty many patients had accessing routine health care. Change in practice, coping with staff shortages and with less face-to-face time with patients caused a lot of general and ethical stress for nurses; nurses felt patient care was compromised.

*“My father is having weekly radiotherapy and chemotherapy which has been missed because different staff give him different instructions as to what he needs to do and no one seems to know or care. One nurse told him he needed a COVID test before coming in for treatment which he had to book, wait for the test, (often the next day) then find a way to get back to the testing clinic, then wait a day for the results. He can’t walk far and has no energy and is now scared he has lost valuable months of life” ID T3203*

As time progressed together with low COVID-19 case numbers seen in Tasmania, nurses were finding the impact of COVID-19 less compared with impact of other personal stresses. Tensions between partners increased as their daily routines changed, personal space decreased and financial pressures increased. Relationships were described as ‘unhappy,’ ‘yelling,’ ‘frightened and scared,’ ‘bullied,’ ‘unpredictable,’ ‘drinking a lot’.

*‘…we already have relationship problems and now with significantly less personal space I feel scared and stressed*.*” ID T1327*

*“I have previously struggled with anxiety & depression, COVID adds another layer. Other stressors in my life like study are also contributing to my general wellbeing. How I feel each day is different, things were definitely worse and more stressful earlier. Now systems are in place and there is planning/ resources it is much better*.*” T3:240*

*“it is difficult to separate feelings and life impacts of COVID and other personal circumstances when they all happened at the same time…….was a tumultuous time in my personal life including divorce, homelessness and job stress. The COVID situation itself actually had very little impact on me in the larger scheme of things*.*” ID: T312*

#### Theme 3. LACK OF SOCIAL SUPPORTS, ISOLATION AND DISCONNECT

With the work environment rapidly changing, so was the nurses’ social environment. Like the rest of the community, nurses faced personal lifestyle struggles. These included home schooling, caring for children with disabilities, managing stepchildren with dual custody, providing ongoing support to elderly relatives, the impact of family unemployment and the change to online tertiary education. Nurses were always cognisant of the potential of viral transmission from the workplace to the home. Nurses were self-isolating from family to prevent transmission from work. Nurses commented at length about the isolation from family locally, interstate and around the world, from saying goodbye to dying relatives and attending funerals.

*“4 kids at home-several weeks of home schooling (before school holidays) on days off-huge learning differences between kindergarten and Gr 9. Chaotic house…. Both of us essential workers (also picking up additional hours to support the COVID demands) but some fear regarding keeping kids at school when advice is to keep home if you can…. No family in south, they are in NW where COVID outbreak occurred!……..elderly neighbours in quarantine from cruise ship so shopping for them as well. Lot of products unavailable and quantity restrictions in supermarkets is stressful for large family, in addition to queuing, packing own bags etc. Studying COVID every night and weekends, ever changing information and advice. Trying to disseminate information and make changes at work accordingly…..Poor internet reliability, not enough technology for all members of family to be schooling on-line at once, data insufficient for additional use at home. Updating Will. Not enough hours in the day to get everything done resulting in less sleep*.*” ID*: T256

*“I care for my aged mother who lives on her own with dementia. The support groups are no longer able to provide the same amount of care for her which has added more pressure on family members…” ID: T2299*

*“My parents living in London and I don’t know if I will see them again*.*” ID: T3450*

*“I decided to self-isolate and live at our holiday home to protect my family*.*” T2:52*

#### Theme 4. FINANCIAL AND ECONOMIC IMPACT

While the COVID-19 pandemic caused much financial hardship and economic downturn throughout the nation with effects felt worldwide, most study respondents felt relatively unaffected as essential workers, grateful to have employment. Many indicated they saved money due to free childcare entitlements, less discretionary spending, and a halt in face-to-face allied healthcare spending. Still, nurses endured the effects of widespread unemployment of family members or family business and superannuation investment losses. Loss of the second incomes resulted in mortgages put on hold.

*“We (my husband and I) have been fortunate enough to work in a low-risk area in a position that keeps requiring us to be at work. This is fortunate because our children may need our assistance in maintaining a roof over their heads*.” T3:82

*“Have jobs!! Very lucky to work in essential services. Even picking up additional hours to help out with COVID response. Superannuation impacted. Kids fees for after school activities cancelled despite paying term fees. Cheaper petrol a huge bonus! Parking for work less stressful with less traffic and demands for parking. Youngest child in childcare one day/week-now free for a term or so if I get her back in Term 2, this is definitely a bonus. Some businesses offering discounts to essential workers-enables you to utilise them more readily and bypass supermarket occasionally*.” T2:374

Nurses facing immediate retirement indicated the loss associated with superannuation investments would impact their retirement plans by extending their working life.

*“My superannuation has taken a hit and I will have to work longer*.*”* T2:62

Changes to pandemic related leave entitlements occurred initially causing confusion with the lack of clarity around leave while waiting for test results. Annual leave and long service leave entitlements were used to supplement sick leave.

*“Having to take sick days and get tested every time I get a cold, once negative there is pressure to return to work with staff shortages despite potentially being infectious still as virus’ are more than a 2-day thing. Sick leave has run out, have a very vulnerable cohort of patients too*.*” ID*: T194

#### Theme 5. WORK-RELATED PRESSURES

The chronic shortage of nurses was compounded as numbers dropped further. Health services immediately changed their model in an attempt to re-direct resources to the emergency pandemic cause.

*“the instability of each day has been the hardest thing to work through, change and information fatigue is the thing I am feeling most. My ability to keep my ‘day to day’ work on track has been impacted greatly*.*” T2: 79*

However, absenteeism of health care staff placed additional demands on the remaining workers, leading to increased workload, unsafe staffing levels and in turn increased sick leave. Staff leave was cancelled. Nurses indicated the work stress was causing sleep issues, with many concerned about the plight of colleagues intrastate and interstate.

*“The thing that has impacted me most at work is that others are not coping - huge impact on absenteeism across the workforce*.*”T3: 289*

*“It was really hard as some nurses were coming to work but lots weren’t. This meant we worked down. Yesterday on the late shift, we normally run with 11 nurses, we had only 5 and 3 of them were doing doubles. I had 6 patients needing full hoists that day. Families get angry with us for not giving their loved one enough care. This happens day after day and we’re getting so tired*.*” T1:451*

Increasing capacity meant changes to hospital infrastructure seeing the relocation and redesign of acute wards and services. Increasing human resources is always a priority in nursing and April 2020 saw the commencement of nursing graduates to the workforce, adding to the stress of existing staff and the transition of graduates very challenging.

*“Approx 130 Graduate nurses have started, how are these already stressed young nurses at the start of their career meant to feel supported, when the current staff feel the ongoing stressors and struggle to come to work?” T3:561*

*“I’m mature aged and a late starter in nursing. One of the most stressful careers I have ever had and not sustainable*.*” T2:29*

*“I’m much more disengaged with the workplace than ever before now. I seek fulfillment in other areas so nursing is now just money” T2:413*

Many nurses were forced to change and expand their role and responsibilities. As intensive care departments were put under pressure, staff from other areas were told they may be redeployed to assist in these areas. These ‘surge’ nurses described training as rushed, stressful and inadequate, seeing staff from acute and subacute areas completing short online education with no associated clinical training. This left experienced intensive care nurses feeling devalued, fearing standards of patient care would not be maintained and raised the issue of building capacity for critical care nurses in the future.

*“This training devalues all the experienced critical care nurses*.*…I only have 3 years of experience and still feel junior…it takes years and years of experience to work here” “T3:455*

Overall nurses felt there was a lack of acknowledgement and understanding of the impact of COVID-19 on their role change by workplace management. Nurses described a department culture of yelling, belittling, passive aggression resulting in low job satisfaction. Nurses described tense relationships with management with little recognition of staff.

*“…we are always pushed to do more and more; management does not acknowledge how tired and stressed staff are*.*” ID: T2314*

*“The times I feel anxious are the hours before I go to work and when team leaders are not carrying out instructions as they should be. I try not to let them get to me, but anxious staff are making me feel more anxious, particularly when they are supposed to be more experienced than me! It’s hard putting on a brave face all the time as it is exhausting*.*” ID: T3266*

*“I think I would have liked regular debriefing and problem-solving sessions on a ward level (more difficult due to social distancing etc. We eventually set up Facebook group, but then had to set up another one as our temporary manager at the time did not like us venting our frustrations). There was an online stress reduction program but finding time to do that after work was not realistic and found it did not address specific concerns I had. Mostly about how do I manage my home life and be a nurse at the same time and keep everyone safe.… “ ID: T3269*

#### Theme 6. POSITIVITY AND RECOVERY

Nurses wrote at length about the positives of the COVID-19 pandemic. They described positive benefits, particularly in home and family relationships, more time to focus on reading and gardening and valuing their job security. Nurses never lost sight of the difficulties their colleagues were enduring locally and internationally and considered themselves very grateful to have a lifestyle of relative freedom and economic and health security.

*“I reduced my work hours and undertook value reflection as a result of COVID. My quality of live and feelings of happiness have improved as a result*.*” ID: T3321*

*“We are more financially secure now and more connected as a family than ever. Home-schooling works for us, our mortgage is on hold for 6 months and my partner is currently being supported with a Jobseeker payment. Applying for Jobseeker was stressful and before commencing home school we were anxious about how to do it, but now these things are in place we are not stressed)*.*”ID: T3466*

By 12-month timepoint, nurses were working towards the COVID-19 vaccine roll out which provided a focus of positivity and end to the pandemic.

*“I feel like I’m getting through the worst of it now! I’m normally anxious & going through perimenopause, so i don’t think COVID can be blamed for everything! Also, it feels like the THS [Tasmanian Health Service] is a little more informed & ready now, so that’s comforting*.*” ID: T3271*

## DISCUSSION

While the COVID-19 pandemic was foreseen by the Commission on Global Health Risk (49) in 2015, governments did not invest in pandemic preparedness and as predicted, the delay cost the world many lives and livelihoods. As the largest cohort of health professionals, nurses and midwives around the world were thrust into the public eye as they waged the war against COVID-19. Health service management have a responsibility for the welfare of its staff and optimum service delivery.

This study was accessible by all public sector nurses and midwives in Tasmania, with 66% of study respondents greater than 51 years, which is an older representative sample compared with the average age of nurses in Australia of 44.3 years (50). Response rates changed over time with notable drop-off at the 3- and 12-month timepoints. The survey was promoted at different times across the three Tasmanian regions due to the changes occurring in response to the pandemic according to regional needs. This variation in the timing of survey distribution may have had impact on response rates, together with survey fatigue; witnessing a plethora of staff surveys circulated at this time. With fewer COVID-19 cases in Tasmania (Tasmania registering only 7 new cases of COVID-19 in the 12 months up to May 2021 (51) nurses may have seen the survey as a lesser priority than in the initial months.

The immediate area of concern for nurses and midwives as the pandemic unfolded was disease transmission prevention and mitigating spread. The focus quickly became personal protection; the correct, efficient, and effective use of PPE. Nurses and midwives articulated PPE as a source of stress: access to PPE and the correct use of PPE, which is consistent across the literature (21). Nurses and midwives concern about adequate access to PPE was reflected across all psychological domains, showing as concern about adequate access to PPE increased, so did levels of anxiety, depression, and insomnia. Stress associated with potential COVID-19 exposure was high and was coupled with emotional exhaustion (high levels of stress, anxiety, depression aligning with rates of insomnia). The sudden reliance on PPE by nurses who do not regularly use PPE, the abolition of fit testing (except in high-risk areas) and the introduction of the staff PPE ‘buddy’ system to support precautionary measures all signalled change, and sources of anxiety. The organisation’s strategy to provide a PPE training video for consistent education and training became both a stressor and support. It was supportive being available on demand, however a stressor for those where PPE use was unfamiliar. The introduction of PPE buddies exposed knowledge and practice deficits among staff who regularly use PPE, leaving staff feeling scrutinised. Working in wards, corridors or medication/treatment rooms and sharing staff tea rooms and communal areas where reducing staff density was not feasible, represented double standards for nurses and heightened their risk of exposure and transmission. The use of PPE for other healthcare associated infections were reduced so PPE could be conserved for COVID-19 patients. These eroded the confidence in infection control systems and the perceived organisational support.

Working in PPE is hot, physically restrictive, makes communication difficult and is both physically and mentally exhausting. While actual numbers of COVID-19 cases were low in Tasmania, there remained a constant stream of suspected COVID-19 admissions throughout Tasmanian hospitals as the state supported repatriation of international and interstate residents, essential workers including seasonal workers and border re-openings. Nurses and midwives working in COVID-19 designated areas were affected by the constancy of the increased workload, and constant threat of COVID-19. Free text comments uncovered this group of nurses and midwives who felt isolated and unrecognised, suffering mental stress and exhaustion while the majority felt all was ‘back to normal.’ The overall psychological effects of stress and mental exhaustion related to PPE were seen to slightly diminish by the 12-month timepoint, yet there remained a minority of nurses still working in the high-risk zones throughout Tasmanian hospitals, suffering mental stress and exhaustion, which study numbers do not readily reflect yet are evidenced in free text responses.

Levels of stress, anxiety, depression, and insomnia relating to the current family and home situation were high with levels continuing to climb throughout all surveys. Respondents free text spoke about the impact of the pandemic on their personal and family lives. This study shows nurses and midwives shared similar fears to the general population. They feared the unknown, feared for their family and significant others which became heightened with border closures, preventing many accesses to their friends and family’s interstate and internationally for long periods. Many were impacted by family bereavement, unable to attend funerals or celebrate family events. This effect was evident at all time points as the strain of lack of contact continued to be felt, triggering a process of grieving and a sense of loss of missed time and opportunities. Many nurses felt at risk, stating ‘they did not sign up for this’ describing unacceptable levels of risk for healthcare professionals during the pandemic. Nurses and midwives described their discord towards Government power, nature of consent and human rights when it came to proposed mandatory vaccination for healthcare workers all the while protective of the capacity of the healthcare system and society.

Financial stress soared to high levels among respondents, peaking at 3 months though remaining at extremely high levels throughout the 12 months. Results show many nurses and midwives and their families suffered financial hardship and were very grateful to have assured employment while others felt financially stronger though economically vulnerable.

Lack of clear communications and advice greatly contributed to the stress felt by responders as they attempted to make sense of their rapidly changing world. ‘Fear spreads even more quickly than infections,’(49) particularly true up to the 3-month timepoint. Nurses and midwives indicated extremely high levels of stress and anxiety in relation to the inadequacy of information and the misinformation generated from both the general and social media. During this time and equipped with very little pandemic preparedness, the organisation was forced to undertake considerable role change which included the formation of the COVID-19 emergency-response management structure. This strategy unsurprisingly resulted in confusion, unclear lines of responsibility, accountability and authority for hospital executives and senior clinicians. Initially information flow seemed blocked; strategic plans made at senior management level were not translated to staff on the frontline. Nurses and midwives were chastised for not following directions by senior management however were not aware of the strategic plan. By 12-month timepoint ward relocations had been completed, systems of communication were now structured and becoming embedded, and health care workers were understanding and adapting to the overall coordination of the pandemic. Measured levels of stress were seen to reduce by 12-months timepoint.

Past studies looking at nursing culture (52) show nurses place great value in workplace team support and the pandemic has shown two extremes; both nurses and midwives working together and the other extreme of feeling alone, isolated, and unsupported. Maintaining workplace team support was of great concern for nurses, with results showing extremely high levels of anxiety, depression, insomnia and general stress relating to concern for adequate team support increasing across the study period. Workplace team support has been shown to have direct effect on patient safety and missed care (53) and burnout (54). Given the downturn of the perceived threat of COVID-19 at the 12-month timepoint yet persistently high levels of distress with an increasing trajectory across all domains, highlights the need for further research to understand the ongoing causes of stress and mental exhaustion in the organisational domain. Australian Institute of Health and Welfare acknowledge burnout as part of the 11th Revision of the International Classification of Diseases (ICD-11-AM) (55) as an occupational phenomenon resulting from chronic workplace stress, not successfully managed. Study data show risk factors associated with a pre-disposition to burnout.

Respondents’ comments indicate exhaustion stemming from working long hours with unrelenting pressure which has seen to precipitate both absenteeism (56) and presenteeism (57) due to high mental distress rather than physical unwellness. Nurses’ often put the needs of the team before their own and hold a cultural ideal of team loyalty (58). Nurses will come to work when they are sick to not let the team down. While COVID-19 screening for respiratory illness may have reduced the rate of presenteeism from physical unwellness, (59) study results show there were high levels of presenteeism among Tasmanian nurses suffering mental distress.

As a profession, nurses are altruistic (60) having a strong sense of doing the best they can with limited resources available (61). Study results align with this trait, showing nurses carry high levels of stress, anxiety, depression and stress in relation to the provision of poor-quality care, particularly up to the 3-month timepoint. We saw nurses still going to work, under greater pressure with the desire to do ‘what was right’ for the patients; struggling to meet their own high expectations of quality patient care yet unwittingly subjecting the patients to high risk of adverse patient safety events (62). At times the sense of not being able to do enough was heightened when nurses felt alone and at times in a professional void without an end in sight. Free text comments show the words ‘burnt out’, ‘disengaged’ with nurses questioning their loyalty for the profession into the future. Nevertheless, the additional demands placed on nurses during the pandemic were not conceptual, they were very real. For nurses, the imperative of providing high-quality patient care is very highly valued, integral to their professionalism. The responsibility for balancing quality of care with availability of resources rests firmly with senior political and professional managers of the health service. In practical terms, for nurses, there was little recognition of that responsibility, and no explanation of how they might be going about managing this balance. As a result, the pressure-points were transferred to the frontline workers, who were unable to handle the balancing or the strategic planning processes. At the 12-month timepoint, levels of concern start to reduce though study results cannot confirm this trend and recommends further study this area.

Health services normally operate in a climate where there is always more demand than can be satisfied. The COVID-19 pandemic placed massive additional demand on nurses and midwives particularly in public hospitals, taking their pre-existing high stress to extreme levels or overload. Health service management responded by cutting-back the provision of other services, however this resulted in the move from familiar practice to unfamiliar practice. To a variable extent, management was aware of this and has responded with varying degrees of skill and comprehension.

By 12-months, surge plans and capacity preparedness became part of the norm. The World Health Organisation provided guidelines for organisations (27), reinforcing that as the pandemic progressed, more information and scientific knowledge will be gained, and the response enacted accordingly. Among emergency service operations and planning, the question arises as to the adequacy of plans with respect to dealing with the psychological effect of demand changes, and whether the line managers were adequately trained and appropriately selected to implement the plans that were devised. Even if the emergency plans were appropriately designed, the question remained around what to do about the care burden of postponed services, and the nagging awareness of that deficit in service among the health professionals who were required to change priorities.

High personal expectations enhanced by a sense of duty, coupled with professional and personal demands placed on nurses by the pandemic created personal discord. It was evident at the 3-month timepoint nurses were being confronted with anti-social public behaviour and recommended to wear plain clothes when travelling to and from work to minimise the risk of abuse. Such behaviour placed even more stress on nurses, the profession who are protectors, now requiring protecting and measures to look after themselves in order to care for the patients. Though nursing and midwifery are stressful professions (63) stress may originate from the working environment, individual or organisational factors or a combination.

By 12-month timepoint, plans for the COVID-19 vaccine roll out had commenced. Nurses were able to access the vaccine as frontline workers, and this began to give some a sense of relief that there would be a slow return of border openings and with that national and international economic stability, reducing uncertainty and frustration.

This study was very sensitive to the possibility questions may precipitate existing stress among respondents. Levels of extreme stress with thoughts of self-harm and feeling as though there was no end and no way out were uncovered in study responses. Extreme levels of stress have the potential to manifest as PTSD which need to be addressed by organisations. The study survey provided links to support services for respondents.

### Study Limitations

Study limitations include the broad classification of nurses without providing the breakdown of nursing positions and therefore cannot accurately describe the stress associated with nursing positions.

Study authors underestimated the longevity of the pandemic. Extending the study beyond one year would provide a truer reflection of the long-term psychological health of nurses and its association with the pandemic or association with pre-existing organisational factors.

## CONCLUSION

Entering a pandemic unprepared, proved a test for the community, health system and esprit de corps. Primary findings include:

Workplace team support directly affected stress and mental exhaustion among nurses and midwives and a strategic priority should include building and strengthening the workplace team, supports and culture into the future. The provision of quality patient care shows a direct effect on levels of stress and mental exhaustion with the lack of quality patient care causing staff disengagement. Nurses described ethical stress from practice change; forced to adopt practices which did not adhere to best practice due to social distancing (such as changes to clinical handover, ward rounds and team meetings, meal breaks, management, and committee meetings). Strategies looking at resource allocation against the provision of quality patient care are essential for the mitigation of adverse patient risks may assist in nurse retention.

The impact of COVID-19 on current and future nurses and midwife’s family, home, financial and economic domains, caused extremely high levels of stress and mental exhaustion. While the provision of stable and functional relationships at home cannot be controlled by the health service, there may be opportunities to provide targeted support (e.g. in domestic violence situations). Understanding strategies to provide support at home for nurses and midwives may provide protection, social care, improve wellbeing and the culture of valuing nurses and midwives.

Lack of a strong communication program proved an important factor for nurses and midwives during the COVID-19 pandemic. The initial 3-month period of COVID-19 was particularly difficult for nurses and midwives, dealing with the rapidly changing scene with no clear governance. Provision of up-to-date information around workplace processes and patient access including staff and patient safety are paramount. Professional employment issues, such as pandemic leave and associated changes to leave entitlements were also rapidly changing leaving nurses and midwives without clear scope of employment boundaries. The provision of clear and specific information and processes for health care workers may help to mitigate this confusion and stress in the future.

Issues relating to PPE was the focus of much stress and mental exhaustion. Nurse’s regard being protected at work as a priority for themselves and their families and felt this reflected the level respect and regard the organisation had for their profession. Furthermore, the overall provision of PPE management and the idea of “just-in-time” inventory management has been shown to be a serious mistake in any event that have major disruptive effects on supply-chains. The safety of all healthcare workers must be a priority for the organisation, ensuring readily available and accessible stocks with staff being trained in its use, in the future.

This study shows younger, female nurses and midwives suffered higher levels of stress and mental exhaustion within the cohort. Planning projections for the nursing workforce show that Australia’s demand for nurses will significantly exceed supply, with a significant projected shortfall in total nursing workforce by 2030 (64) due to an aging workforce. Teaching resilience will help support the future nursing and midwifery workforce rather than fuel further attrition while fostering retention.

Recommendations for managers, before the memories of COVID-19 fade:

A health service is reliant upon its staff to deliver its health services. It has an imperative to deliver high quality management, firstly to ensure optimum delivery of care and secondly to assure the welfare of its staff, despite the inevitable health stress imposed by the nature of the work.

1. Workplace team support is a critical driver among nurses and midwives and if not met can precipitate stress and possibly burnout. Burnout is recognised in the ICD-11-AM and systems to monitor burnout among nurses and midwives need to embed within the organisation’s culture.
2. The provision of quality patient care is a critical driver for organisational culture among nurses and midwives. Inclusive strategies looking at resource provision against optimal service delivery will potentially strengthen organisational culture for nurses and midwives, mitigate adverse patient risks and assist in nurse retention.
3. The design of a strong program for communication with processes specific for health care workers using prompt alerts and rapid dissemination of relevant information are essential for effective outbreak response.
4. The safety of all healthcare workers must be a priority for the organisation, with readily available and accessible PPE stocks with staff being well trained in its use.
5. Building resilience particularly among the younger members of the nursing and midwife team will help preserve the mental health of the upcoming nursing workforce, protecting against further attrition and potentially increase retention.
6. Provision of family, home, and financial support for nurses and midwives will protect the health workforce and the provision of quality patient care. While the health service cannot directly control the relationships at home, there may be opportunities to provide targeted support.

### Recommendations for further research

Trial systems to monitor burnout among nurses and midwives within the healthcare organisation Trial interventions to support nurses and midwives which foster resilience.

## Data Availability

All data produced in the present study are available upon reasonable request to the authors

## Author Contributions

Conceptualization, methodology KMM & JP; formal data analysis and reporting, IKR & KMM; data curation, IKR & KMM; data interpretation, KMM, JP, IKR; writing original draft preparation, KMM; editing input and review, KMM, JP, IKR; supervision and project management and administration, KMM & JP; All authors have read and agreed to the published version of the manuscript.

## Institutional Review Board Statement

The study was conducted according to the guidelines of the Declaration of Helsinki and approved by the Human Research Ethics Committee (Tasmanian Network) Reference No. H0021677 on the 29^th^ April 2020.

## Informed Consent Statement

In line with the Tasmanian PIP Act 2004 defines sensitive information. This is a low-risk on-line survey of nurses and midwives in Tasmania. All data accessed was de-identified and coded prior to exporting for statistical analysis to protect confidentiality. Any reported data has been pooled amalgamated data with no identifying features. Safeguards are in place for appropriate and ethical use of data and limited access by nominated researchers. The data is appropriately archived to protect confidentiality. Waiver of consent was sort and approved for the ethical use of the data prior to commencing data collection.

## Data Availability Statement

Authors will supply data upon request.

## Acknowledgments

The authors gratefully acknowledge the Clifford Craig Foundation, the University of Tasmania and the Tasmanian Health Service for their support in kind, providing statistical analysis and input from all investigators to develop and support this process.

